# A randomised control trial of the effects of home-based online attention training and working memory training on cognition and everyday function in a community stroke sample

**DOI:** 10.1101/2021.04.01.21254686

**Authors:** Polly V. Peers, Sarah F. Punton, Fionnuala C. Murphy, Peter Watson, Andrew Bateman, John Duncan, Duncan E. Astle, Adam Hampshire, Tom Manly

**Author notes:** Corresponding Author: Polly Peers, MRC Cognition & Brain Sciences Unit, 15 Chaucer Road, Cambridge, CB2 7EF, UK.

## Abstract

Cognitive difficulties are common following stroke and can have widespread impacts on everyday functioning. Technological advances offer the possibility of individualised cognitive training for patients at home, potentially providing a low-cost, low-intensity adjunct to rehabilitation services. Using this approach, we have previously demonstrated post-training improvements in attention and everyday functioning in fronto-parietal stroke patients. Here we examine whether these benefits are observed more broadly in a community stroke sample. Patients were randomised to either 4 weeks of online adaptive attention training (SAT), working memory training (WMT) or waitlist (WL). Cognitive and everyday function measures were collected before and after intervention, and after 3months. During training, weekly measures of patients’ subjective functioning were collected. The training was well received and compliance good. No differences in standardised cognitive tests were observed for either intervention relative to controls. However, on patient reported outcomes, SAT participants showed greater levels of improvement in everyday functioning than WMT or WL participants. In line with our previous work, everyday functioning improvements were greatest for patients with spatial impairments and those who received SAT training. Whether attention training can be recommended for stroke survivors depends on whether cognitive test performance or everyday functioning is considered more relevant.

## Introduction

Stroke is the single biggest cause of long-term disability in industrialised nations. Post-stroke impairments in attention are common, debilitating and associated with slowed recovery and increased reliance on public services (Barker-Collo, Feigin, Parag, & Lawes, 2010; Hyndman & Ashburn, 2003). Surveys of stroke survivors indicate that difficulties in attention and concentration are rated as among the most problematic persistent symptoms (Stroke Association, 2018 https://www.stroke.org.uk/h4B).

A distinction is often drawn between impaired *lateralised spatial attention*, in which patients can have sometimes striking difficulty in noticing information on one side of space, and *more general problems* in filtering relevant from irrelevant information and maintaining an alert, engaged, goal directed state. Problems with spatial attention (variously termed *Unilateral Spatial Neglect, Hemi-inattention, Hemi-neglect*) have a well-established relationship with slowed recovery, heightened dependence on others and problems with everyday activities (Jehkonen, Laihosalo, & Keetuenen, 2006; Katz, Hartman-Maeir, Ring, & Sorojer, 1999). Accordingly, there has been considerable work proposing and evaluating a wide variety of methods to reduce spatial bias. Logically enough, most of these techniques have aimed to directly correct spatial bias, for example, by training eye- and head-movements to the neglected side (Luukainen-Markkula, Tarkka, Pitkänen, Sivenius, & Hämäläinen, 2009), or by inducing such movements via adaptation to prism lenses (Rossetti et al., 1998), or hemifield patching (e.g., Rossi, Kheyfets, & Reding, 1990). Whilst all of these methods have had demonstrable impacts on bias as measured in sensitive tests, there remains insufficient evidence of generalised and long-term benefits on everyday activities (Bowen et al., 2013). Whilst the lateralised impairment of spatial bias seems distinct, there is growing evidence of a relationship between it and more general, non-spatially specific problems in sustaining attention and maintaining an alert state. Spatial bias is more likely to arise and persist in the context of these more general attentional impairments (Duncan et al., 1999, Peers et al., 2005, Habekost & Rostrup, 2006; Corbetta and Shulman 2002, Robertson et al., 1997) and following damage to the ‘non-spatial’ ventral attention network (Corbetta and Shulman, 2011). In line with this, the degree of spatial bias can be modulated in the short term by changes in overall alertness levels, for example, induced by loud tones, stimulant medication, perceived time pressure and tiredness (Robertson et al., 1998; Gorgoraptis et al., 2012; George et al., 2008, Bareham et al., 2014). It is currently unclear whether these short-term benefits could be turned into practical rehabilitation. One difficulty is that people with poor attention/alertness find it hard, almost by definition, to self-monitor and remember to apply strategies (though see Robertson et al., 1996). Is it possible, however, that frequent, systematic training, over a sufficiently long period, could progressively help shape attentive engagement to the benefit of general *and* spatial attention function?

In this respect we were encouraged by the positive results of a 4-week exploratory trial of daily home-based attention training with precisely these aims (Peers et al., 2020). Specifically, we recruited volunteers from a panel of people with previous acquired brain injury interested in taking part in research. We focused predominantly on those with right fronto-parietal lesions in whom attentional impairments were likely. They were randomised either to home-based attention training (SAT), a similar home-based working memory training (WMT), or to a waitlist control (in which the pre- and post-measures were administered at the same interval but with no intervening training). The training programs were well matched in both using game-like tasks that adaptively increased the challenge as participants’ performance improved (although it should be noted that there were differences in the feedback provided and potentially also in the duration of the two training regimes due to exercise-end rules based on time for SAT, and number of items completed for WMT – variance that has been addressed in the current study). WMT was therefore a conservative control for the general effects of an intervention and scheduled daily cognitive exercise. Engagement, compliance and self-management of the training in both cases was good. As with other reports, we found that WMT was associated with rather specific improvements on an untrained working memory task. SAT, in contrast, was associated with a broader range of improvements in attentional capacity *and* significantly reduced spatial bias. Reductions in spatial bias were, in turn, most predictive of *self-reported* improvements in cognition and everyday activities. The latter is important. There has been quite extensive discussion of the relative value of physiological and behavioural measures, clinician reported outcomes, and patient reported outcomes (PROs), including in stroke (e.g. Price-Haywood et al., 2019). If the purpose of an intervention is ultimately to improve subjective evaluation of quality of life and satisfaction then it is important to measure these features directly rather than assume that they will emerge from, for example, improvements detected on standardised cognitive tests.

These results from Peers et al. (2020) were encouraging but caution is required. There is rightly concern that positive publication bias, in conjunction with small group sizes, inevitably tends to exaggerate therapeutic benefits. If people are to be encouraged to spend even 20 minutes a day training it is important to know; a) whether these results would reliably replicate in a much larger sample, b) whether attention training may be of benefit to the wider stroke survivor community, whether or not they have spatial impairments, and c) whether the positive efficacy, feasibility and acceptability findings in part may have arisen from keen, engaged participants with an established interest in research. Answering these questions was the aim of the current study. Because of the strength of the findings in the previous trial and its apparent effect on self-reported improvement, spatial bias – as measured by the Theory of Visual Attention paradigm (TVA; see methods) – was our primary outcome measure.

An obvious challenge to this primary outcome measure is that the prevalence of spatial bias in stroke survivors is estimated at approximately 30% (with wide variations depending on when and how it is measured; Bowen, McKenna and Tallis, 1999). However, it is plausible that spatial bias, as measured on the sensitive computerised paradigm employed here, is much more common than estimates based on (typically single) administrations of a paper and pencil task would generally imply. It is therefore possible that we could observe more widespread improvements in attentional functions using more sensitive tasks, and, by inference from the previous result, knock-on benefits for self-reported cognitive and everyday function. Whether or not this is the case, standard trial practice requires that we select our single, best-evidenced measure which, given the overlap of training with the previous study, must be spatial bias. Once the issue of the primary outcome measure is decided, more nuanced exploration of the pattern of results across our secondary measures can proceed.

Accordingly, as with the previous trial, we compared patients randomised to either one month of daily SAT, one month of daily WMT or a waitlist control and hypothesised that we would observe a similar pattern of results, namely:

- SAT would again be associated with significant reductions on our primary outcome measure (spatial bias), relative to WMT and Waitlist conditions.
- WMT would be associated with specific improvements on untrained working memory tasks similar to those used in the training, when compared with SAT and Waitlist conditions.
- SAT would be associated with a broader range of improvements on untrained attention tasks, as well as reduced spatial bias, relative to WMT and Waitlist conditions.
- Whilst, as previously, both training conditions would be associated with improved participant self-ratings of cognitive function, mood and motivation relative to Waitlist, SAT would be generally stronger in this respect and, specifically, improvements in spatial attention would be the strongest factor driving these reductions in self-reported disability.
- Participants would generally be able to cope with the technical demands on both types of training, find training tolerable and comply sufficiently with the regime to allow inference on efficacy.

## Methods

### Trial design

This was a randomised control trial with post-assessments and follow-ups carried out by a researcher blind to both the intervention and the pre-intervention scores. Participant IDs were randomised prior to recruitment by a statistician, with the allocation of each individual becoming known to the research team only when a sealed envelope was opened at the end of the initial assessment.

### Participants

One hundred and sixty-five stroke survivors living in the community were referred by therapists at Cambridgeshire and Peterborough Foundation Trust (CPFT n = 117) and Cambridge University Hospitals Trust (CUH, n = 11), the Cambridge Cognitive Neuroscience Research Panel (CCNRP, n = 25), and local stroke charity groups including The Stroke Association and “Pos+Ability” (n = 12). Of these 80 finally randomized into the trial (see Figure 1 for details) Patients had all been discharged from acute care and were between 3 weeks and 25 years post their most recent stroke (mean time since injury= 37.7 months ± 59.4 months).

**Figure 1.**
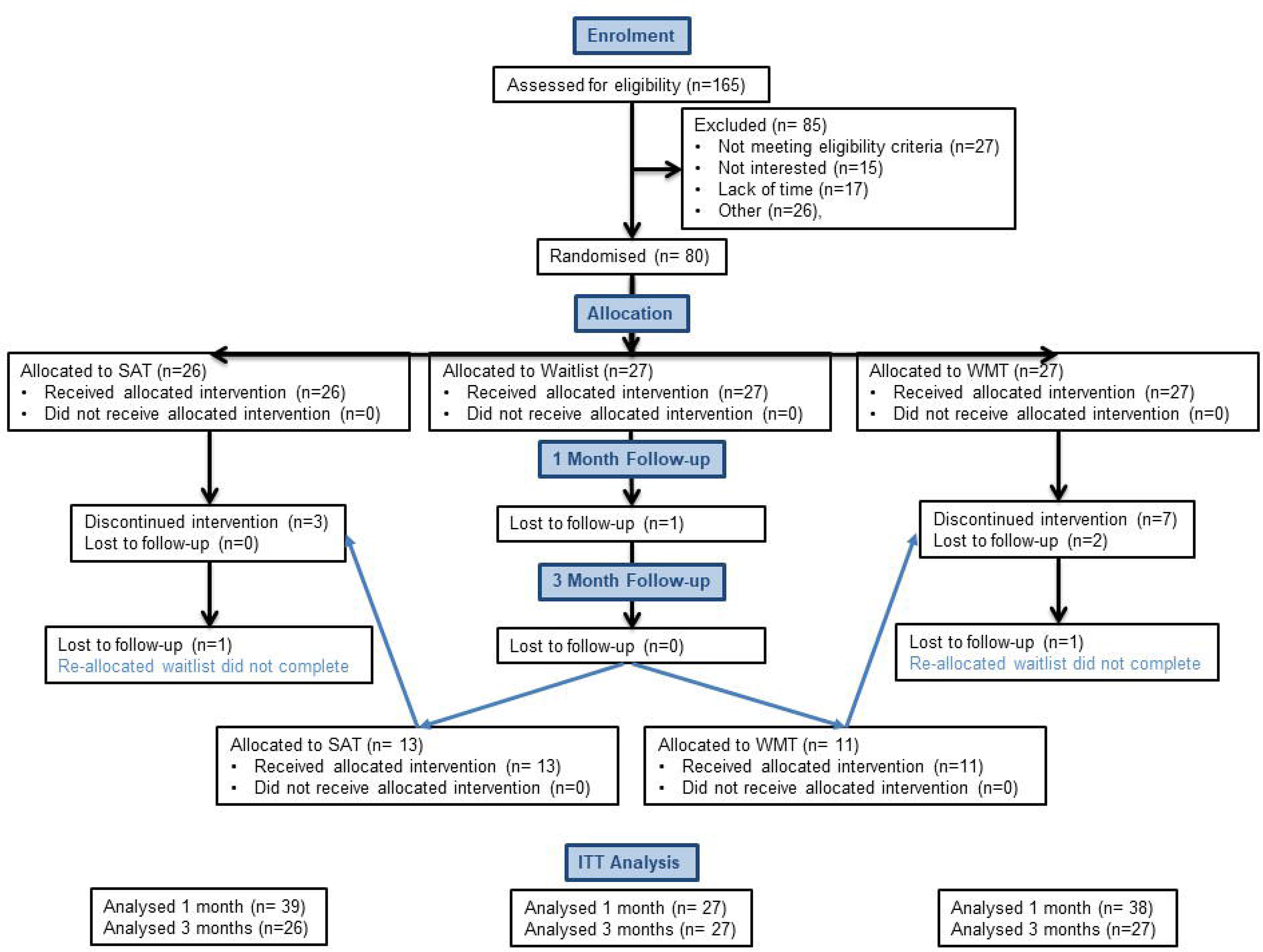
CONSORT diagram showing flow of patients through the study

Inclusion criteria were:: over 18, diagnosis of stroke, proficient in English, and having sufficient language, motor function and general health sufficient to undertake training. In all but a few cases, assessments were conducted in participants’ own homes (otherwise at referring hospital or research unit).

### Interventions

The two active interventions were compared with a waitlist control. Both interventions were developed in-house and had the same basic format and look. Both included a series of 3-5 minute time-limited ‘games’ (4 in WMT and 5 in SAT), designed to challenge the respective cognitive capacities. Both provided trial-by-trial feedback, feedback on overall progress, and incrementally adapted by increasing task difficulty as trainees’ performance improved. SAT tasks typically involved attending to increasing amounts of simultaneously presented on-screen information (e.g. for speeded detection of a target hidden amongst similar distractors) with minimal requirement for holding information ‘in mind’. In contrast, WMT emphasised taking in and recalling incrementally increasing strings of sequentially presented information. In other words, SAT emphasised self-guided deployment, direction and maintenance of attention with minimal memory demand, whilst WMT had high demands on memory but, by typically cueing attention to a single location, had lower requirements for self-directed and maintained attention.

The training sessions took approximately 20 minutes to complete each day, with participants starting the subsequent day’s tasks at the performance level they had achieved at the end of the previous day. Full descriptions and screen shots of the individual tasks can be seen in supplementary materials.

Participants completed the training at home, for the most part on their own computers (computers were loaned in 10 cases). During training/waitlist period, participants were called weekly by a researcher to complete a semi-structured interview covering perception of changes from the previous week in cognitive, mood and social interactions and, where relevant, training duration, perception of training, technical and other barriers encountered. Technical support was provided if necessary.

### Outcomes

Outcome measures were collected in 1-2 sessions at baseline, within 3-weeks of completing training (or 6 weeks of baseline in the waitlist condition) and then again 3 months later. Waitlist participants were re-randomised to a training condition after their 3-month follow-up, with their 3-month follow-up scores acting as the baseline for their intervention. A final set of outcomes were taken within 3 weeks of training completion. Where participants had speech difficulties, point-cards were used to facilitate communication and assessment.

#### Primary outcome measure

Based on our previous findings (Peers et al., 2020), which had shown reductions in TVA whole-report measures of spatial bias as the strongest predictor of self-reported improvements in everyday function, spatial bias was selected as our primary end-point. TVA Whole-report is a computerised assessment in which participants are asked to report as many as possible (6), or a subset (3), of letters very briefly presented in a circular array. This can be used to estimate bias towards preferentially attending to one or other side of the screen as well as visual short-term memory capacity (VSTM; effectively, how much information can be taken in ‘at a glance’). For a more comprehensive description of the tasks see Peers et al., 2020.

### Background measures

The National Adult Reading Test (NART) (Nelson, 1982) was administered to all participants without significant aphasia at baseline as a general measure of premorbid, crystallised intelligence. The Cattell Culture Fair Test was used to measure fluid intelligence (Institute for Personality and Ability Testing, 1973). In addition, visual functions were assessed using the Sloan Letter Near Vision Card (acuity; Good-lite Co, IL) and the Ishihara (1978) Colour-Blindness Test.

#### Secondary outcome measures

##### Attention measures

OCS-BRIDGE is a new, validated touchscreen tablet cognitive and mood screening battery (https://ocs-bridge.com/) that incorporates a range of measures from the Oxford Cognitive Screen (Demeyere et al., 2015). OCS-BRIDGE attention measures used here included: Hearts Cancellation (assessing spatial bias in finding visual targets distributed across the screen amid similar distractors) and Sustained Attention Lateralised Reaction Time (assessing bias in response times to temporally unpredictable changes in brightness of clouds to the left and right of screen, whilst simultaneously counting the number of times a centrally presented lighthouse light flashes).

##### Working Memory Measures

OCS-BRIDGE forward and backward Digit Span tasks and the Automated Working Memory Assessment (AWMA; Alloway, 2007) Dot Matrix and Spatial Span tasks were used to examine participants’ ability to recall and manipulate (e.g. reverse) strings of numbers or spatial locations of increasing length.

##### Measures of disability/symptom severity

The following questionnaire measures of patient and carer perceptions of disability and symptom severity were used:

- The EBIQ (European Brain Injury Questionnaire; Teasdale, 1997): a validated self- and informant-report 63-item questionnaire concerning subjective wellbeing after brain injury.
- The Cognitive Failures Questionnaire (Broadbent et al., 1982): a self-report measure of the frequency of cognitive slips and lapses in attention over the previous week.
- Subjective Neglect Questionnaire (Towle and Lincoln, 1991): an informant-report questionnaire specifically assessing symptoms and consequences of spatial bias.

##### Ratings of symptoms and training experience

A semi-structured interview was carried out each week during training or waitlist. These included participants’ ratings of *change* from the previous week in their memory, concentration, planning, spatial awareness, motivation, mood, social interactions and ‘other’, with values ranging from −2 (“much worse”), through 0 (“about the same”) to 2 (“a lot better”). This formed a cumulative total to track changes across the 4 weeks (e.g. consistent small improvements in an area could give +1, +1 +1, +1 = 4, fluctuations could give +1, −1, +2, −1 = 1 and consistent decline could give −1,−2,−1,−2 = −6; see supplementary materials for details).

Participants completed an additional study completion interview with a researcher in which their views on the study and training conditions were sought (see supplementary materials).

##### Sample Size

A power calculation from our earlier study (Peers et al., 2020) indicated that approximately 33 participants per arm was required to detect significant changes in outcome measures.

##### Randomisation

Two-step randomisation was conducted prior to recruitment by a statistician who supplied participant numbered sealed envelopes, opened by researchers in front of the participant after completing the relevant baseline or final wait-list assessment. In phase 1, participant numbers were allocated to the three conditions (SAT, WMT or WL) using random permutations across variable length blocks (6-15) to achieve approximately equal condition allocation across the trial. In phase 2, participant numbers allocated to waitlist in phase 1 were randomised to one of the two training conditions, here using variable block length sequences of 2-8 participants. Because block length/sequence was not known by the researcher, the likely allocation of a given participant could not be inferred based on the preceding allocations.

## Results

The flow of participants through the study can be seen in the CONSORT diagram (Figure 1, below).

### Recruitment and Compliance

The first participant entered the study on 13/05/2015, and the final follow-up session was completed 28/02/2018. Rates of compliance were good, with 92% of SAT, and 82% of WMT participants continuing with the study to the follow-up sessions. There was no statistically significant difference in compliance between SAT and WMT (Χ^2^ (2,77) = 1.96, p=0.16). SAT participants for whom follow-up assessments were undertaken completed a mean of 19.41/20 training sessions (range 14-20) over an average of 34.6 (17-230) days. The three SAT participants who ‘dropped out’ completed 0, 3 and 9 sessions respectively. WMT participants who undertook follow-up assessments completed on average of 18.65/20 sessions (range 15-20) over a mean of 36.3 (15-86) days. The seven WMT participants who ‘dropped out’ completed between 1 and 4 sessions.

### Baseline Data

The 3 groups did not significantly differ on baseline background measures or symptom severity (see Table 1).

**Table 1.**
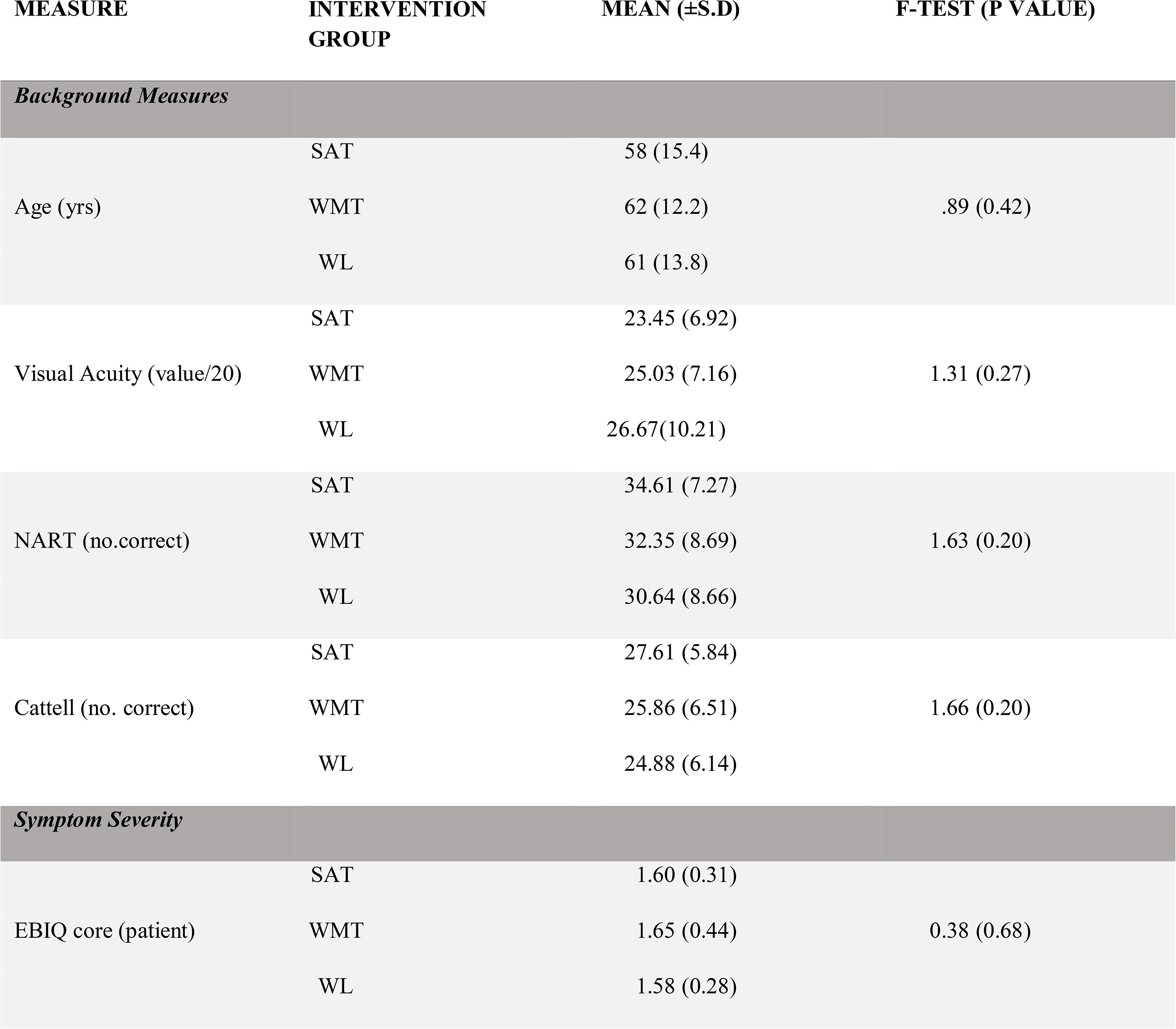
Comparison of the three groups on background measures and self-reported symptom severity at baseline.

| MEASURE | INTERVENTION GROUP | MEAN (±S.D) | F-TEST (P VALUE) |
| --- | --- | --- | --- |
| *Background Measures* |  |  |  |
| Age (yrs) | SAT | 58 (15.4) | .89 (0.42) |
|  | WMT | 62 (12.2) |  |
|  | WL | 61 (13.8) |  |
| Visual Acuity (value/20) | SAT | 23.45 (6.92) | 1.31 (0.27) |
|  | WMT | 25.03 (7.16) |  |
|  | WL | 26.67(10.21) |  |
| NART (no.correct) | SAT | 34.61 (7.27) | 1.63 (0.20) |
|  | WMT | 32.35 (8.69) |  |
|  | WL | 30.64 (8.66) |  |
| Cattell (no. correct) | SAT | 27.61 (5.84) | 1.66 (0.20) |
|  | WMT | 25.86 (6.51) |  |
|  | WL | 24.88 (6.14) |  |
| *Symptom Severity* |  |  |  |
| EBIQ core (patient) | SAT | 1.60 (0.31) | 0.38 (0.68) |
|  | WMT | 1.65 (0.44) |  |
|  | WL | 1.58 (0.28) |  |

#### Levels of impairment

Because there were few inclusion criteria beyond having had a stroke, it is interesting to consider the level of impairments that we observed when participants’ performance on baseline tasks were compared with normative data from the general population. This is shown in table 2 (note: the ‘normative’ data for the TVA task were from a relatively small group (n = 10) of healthy participants who acted as a control for patients in a previous study (Peers et al., 2005) and should be considered as illustrative).

**Table 2.** Participant performance on baseline measures of cognitive function. Here, mean and standard deviations of performance are provided together with the percentage of the sample performing more than 2 standard deviations below the general population mean (i.e. approximately below the 5^th^ percentile), a level often taken as operationalising an ‘impairment’. The percentage of the sample in the less conservative lower 10% range (i.e. below 1.28 standard deviations of the general population mean that may be characterised as ‘probable impairment’) is also presented.

| MEASURE | MEAN<br>(±S.D) | % PATIENTS >2SD<br>BELOW<br>POPULATION MEAN | % PATIENTS >1.28 SD<br>BELOW POPULATION<br>MEAN |
| --- | --- | --- | --- |
| Cattell (scaled IQ score) | 91 (14.4) | 8% | 23% |
| TVA Absolute Bias<sup>1</sup> | 0.09 (0.09) | 35% | 53% |
| TVA VSTM capacity (K) <sup>1</sup> | 3.30 (0.95) | 43% | 65% |
| OCS-BRIDGE Spatial Bias on the Hearts<br>Cancellation Test | 0.063<br>(0.13) | 18% | 52% |
| OCS-BRIDGE Forward Digit Span | 6.05 (1.19) | 8% | 25% |
| OCS-BRIDGE Backwards Digit Span | 4.33 (1.31) | 2% | 49% |
| OCS-BRIDGE Sustained and Lateralised<br>Attention Test (SALT). Spatial bias (omissions) | 0.09 (0.15) | 41% | 64% |
| OCS-BRIDGE SALT Lateralised bias in<br>reaction times | 0.11 (0.24) | 28% | 43% |
| OCS-BRIDGE SALT Sustained Attention | 2.85 (1.02) | 8% | 33% |

Table 2 shows that, if a relatively strict criterion of impairment is imposed, many of the participants showed normal levels of function on challenging measures such as the Cattell Culture Fair fluid IQ and reversed digit span. However, the results from the more liberal 10^th^ percentile criterion suggest that more subtle problems were relatively widespread. Of relevance to our primary outcome measure, the results from the TVA spatial bias parameter and the SALT lateralised reaction time bias measure suggest that the proportion of participants showing significant spatial bias was close to the post-stroke estimated incidence (approx. 30%) reported by Bowen, McKenna and Tallis, (1999). The bias results from the OCS-BRIDGE cancellation measure, which is similar to widely used paper and pencil clinical cancellation tasks lower in terms of strict criterion but, again, broadly consistent when those with ‘probable impairment’ are included.

### Training tasks

As expected, performance on the training tasks improved over the course of the interventions (see Figure 2). Significant improvements in performance were observed in both the SAT, t(35)=-28.50, p<0.001 (mean improvement, 11.01 ±2.32; range 4.07-14.80) and the WMT, t(27)=-9.33, p<0.001 (mean improvement, 1.01±0.62; range −0.04-2.32).

**Figure 2.**
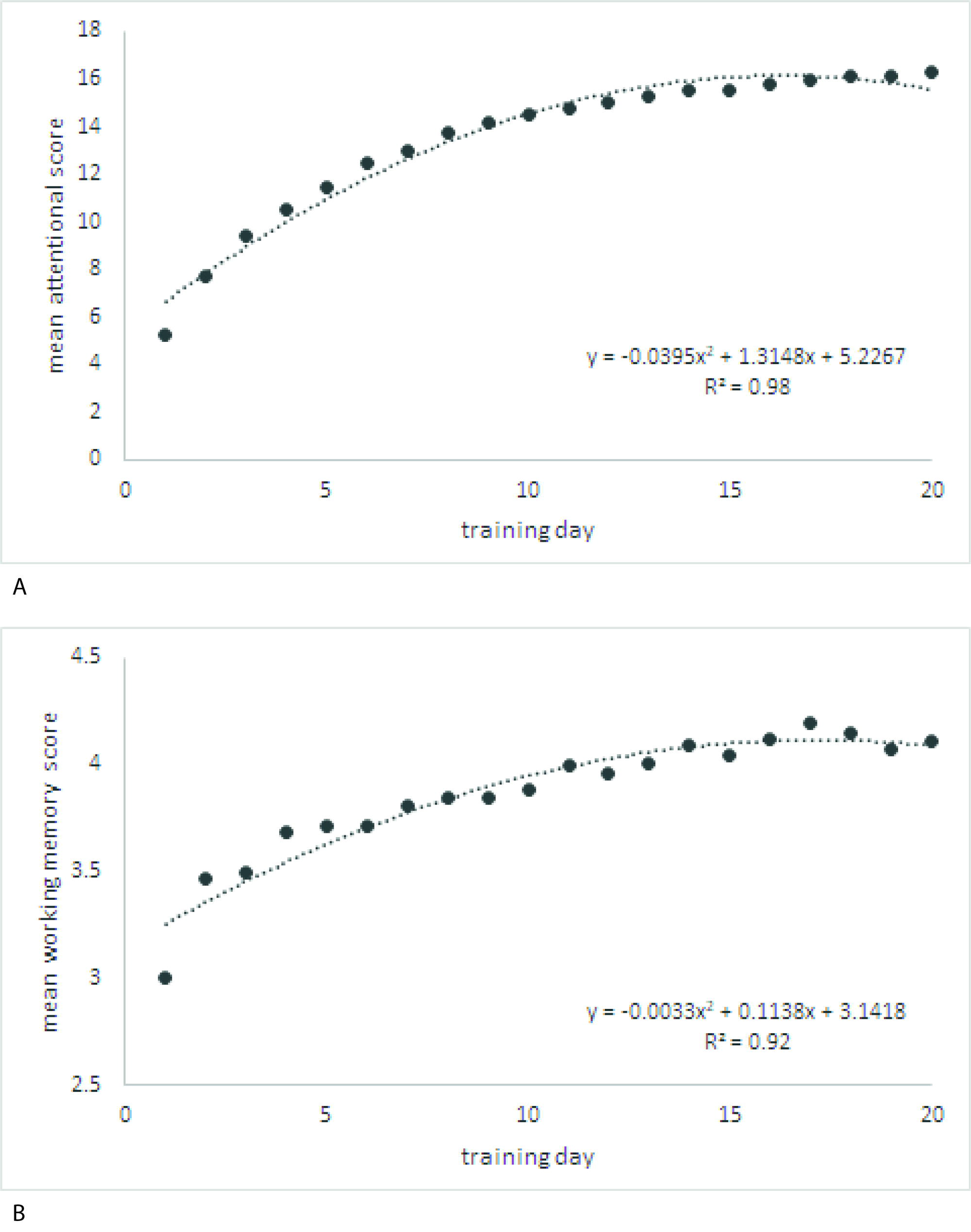
Mean daily performance on the training tasks over the 20 training sessions. A. shows average performance of the SAT patients (collapsed across the five training tasks which made up each session) whilst B. shows the avarage performance of the WMT patients (collapsed across the 4 training tasks they completed at each session).

In the weekly phone calls, participants were asked to rate between 0 (not at all) and 10 (extremely) how helpful and enjoyable they had found the training (see Figure 3). Repeated measures ANOVAs revealed main effects of session (helpfulnesshelpfullness, F(3,192)=36.9, p<0.001; enjoyablenessenjoyableness, F(3, 192)= 9.42, p<0.001); ratings of helpfulness and enjoyableness tended to improve over time for SAT and WMT. There was also a main effect of training type (helpfulness F(1,64)=7.69, p=0.007; enjoyableness, F(1,64)=8.27, p=0.005); SAT participants consistently rated their training as more helpful and enjoyable than WMT participants. There was no group x time interaction; the improvements in ratings over the 4 weeks were not disprortionately greater for either training condition.

**Figure 3.**
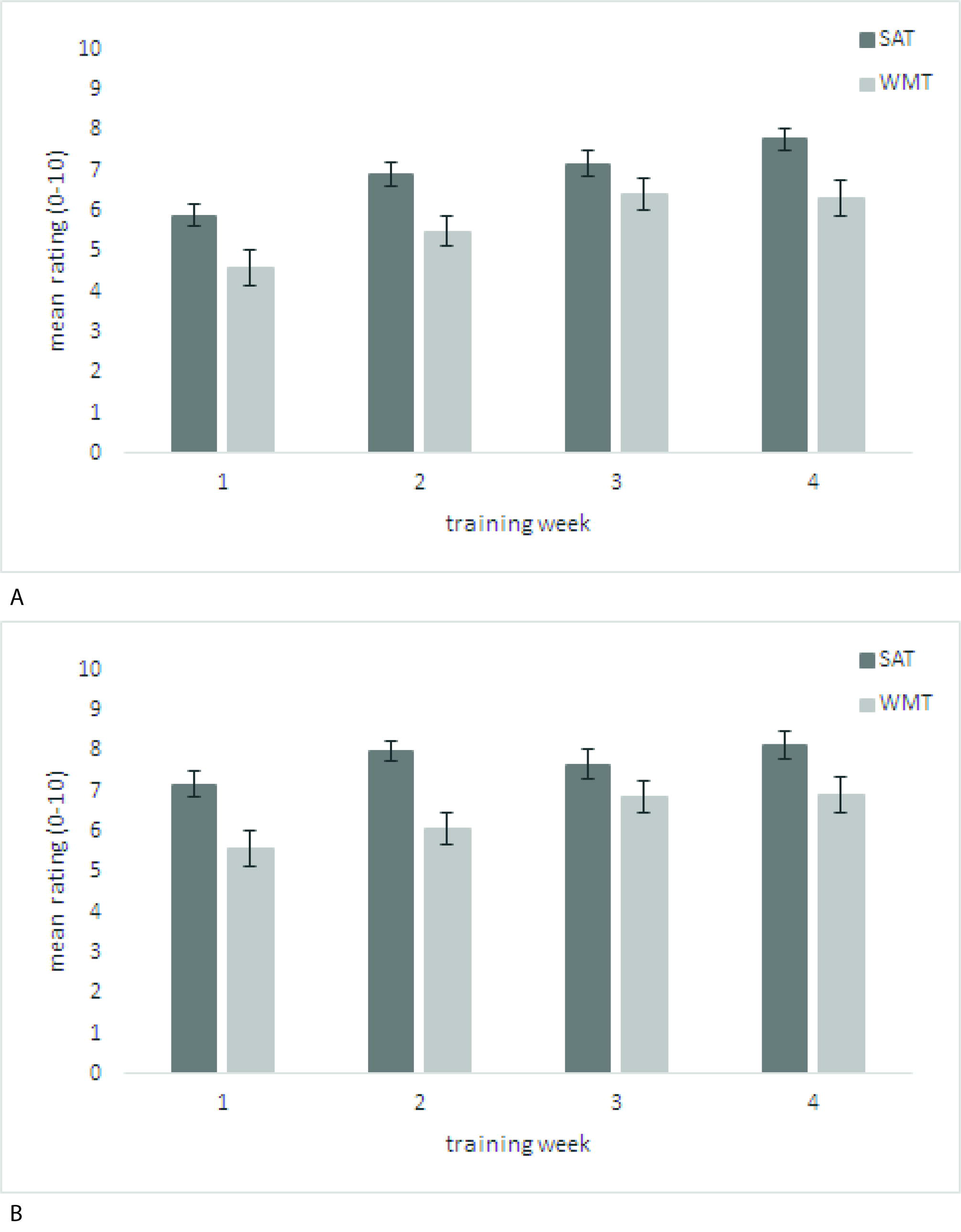
Mean (±S.E) ratings of A. the helpfulness and B. the enjoyableness of SAT and WMT over the 4 weeks of training.

Responses from the study completion interviews also indicated quite high levels of satisfaction with the interventions. Of the SAT participants, 76% thought the intervention had helped them, 15% were unsure and 9% felt that it had not helped. For WMT, 66% felt that it had helped, 17% were unsure, and 17% felt the intervention had not helped. The apparent SAT advantage was not statistically significant (X=1.20, p=0.54). The levels of rated interest and engagement with the training tasks were high for both conditions (SAT 82%, WMT 83%; Χ=0.009, p=0.92). Finally, 100% of SAT and 90% of WMT participants rated their training as manageable in terms of session duration, frequency, technical demands etc.

### Statistical analysis

An enter method regression approach was applied to examine whether post-training/waitlist performance was influenced by intervention group (SAT, WMT ad WL) in a manner that adjusted for pre-training/waitlist performance (for ease of exposition, ‘intervention’ will henceforth be used to refer to the period between pre-and-post assessment, including for the waitlist condition). Coding dummy variables for each group accordingly allowed us to directly compare the effects of the interventions in a single analysis. This regression has been considered a stricter test of intervention gains than the ANOVA approach because interactions in the latter can be driven to varying degree by *pre-*intervention group differences (even if those differences are not, themselves, significant). Where significant regression effects were observed, paired sample t-tests were carried out to examine whether post-intervention scores differed significantly from pre-intervention scores for each of the groups.

### One-month follow-up data

#### Primary end-point-TVA spatial bias

Post-intervention spatial bias scores were significantly predicted (R^2^= 0.67, F (3,97)=64.4, p<0.001) by a combination of ‘pre-intervention’ scores and ‘intervention group’ predictors. ‘Pre-intervention’ score (β=0.82, p<0.001) was found to be the only significant predictor, with neither SAT (β=-0.02, p=0.81) or WMT (β=0.01, p=0.88) groups being significant predictors. The main question of this study was whether improvements in our primary outcome measure (spatial bias) would be disproportionately observed in participants who underwent SAT compared to those who underwent WMT or the waitlist (WL), when those participants were recruited from the general stroke population (i.e. not selected on the basis of attentional impairment)? The answer is a clear no.

#### Secondary end-points

Results of the secondary measures can be seen in Table 3. Much like the primary end-point, the regression analyses for the secondary end-points indicated that post-intervention outcome could be significantly predicted by a combination of ‘pre-intervention score’ and ‘intervention group’. Beta values indicate in all cases ‘pre-intervention score’ significantly predicted ‘post-intervention’ performance, however intervention group did not. Once again, in terms of standardised cognitive measures, the results provide *no* support for recommending either form of training as a general, unselected post-stroke intervention.

**Table 3.**
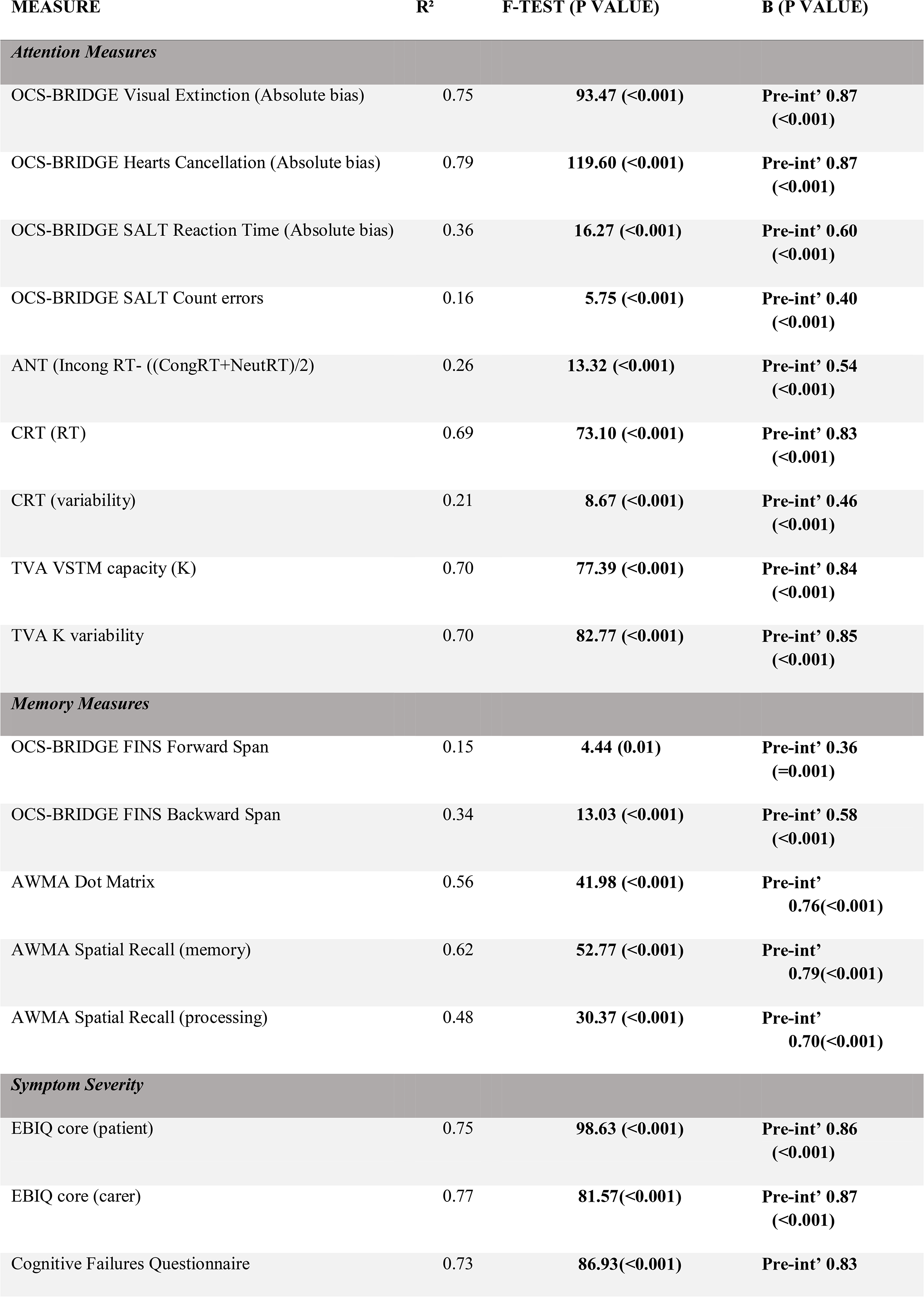
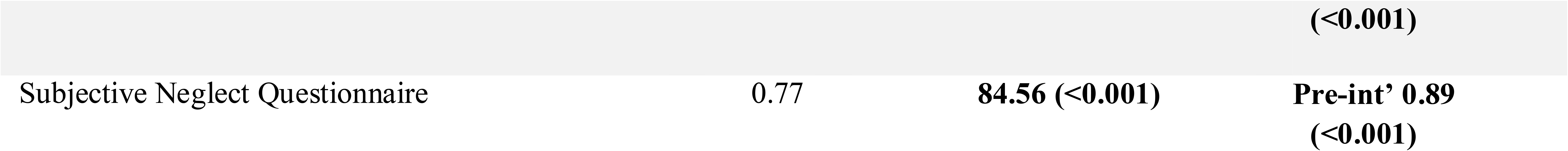
The results of the regression analyses of the secondary end-points. The table provides the variance (R^2^) in post-intervention scores that can be predicted by pre-intervention performance score and intervention group, the F-test and *p* value, as well as the standardised β coefficients (and p-values) of significant predictors. Statistically significant results (P<-0.05) are in bold. Int’ = intervention

#### Self-reported changes in functioning

In addition to post-intervention data, each week participants rated changes in functioning across 8 areas of their lives (memory, concentration, planning, spatial awareness, motivation, mood, social interactions and ‘other’) relative to the previous week. Examining the cumulative pattern of change, collapsed across the 8 domains (i.e. the total at week 4; see figure 4), a repeated measures ANOVA revealed significant effects of time (F(3,303)= 46.40, p<0.001), intervention type (F(2,101)= 7.24, p<0.001), and a significant intervention x time interaction (F(6,303)=12.00, p<0.001). Post-hoc comparisons indicated that the SAT group reported significantly greater improvements in functioning than those in WMT (p<0.01) or WL conditions (p<0.001). WMT and WL did not differ significantly from one another in this respect. To investigate the interaction, one-way ANOVAs were carried out separately on the cumulative totals reached at the end of each week. These indicated no significant difference between the 3 groups, F(2,103)= .89, p=0.41 at week 1, with a significant effect emerging at week 2, (F(2,103)=4.31, p<0.05) that strengthened at weeks 3 (F(2,103)=9.11, p<0.001) and week 4 (F(2,103)= 9.61, p<0.001). Post-hoc tests revealed significant differences between SAT and WL by week 2 (p<0.05), which broaden to significant differences between SAT and both WL and WMT at weeks 3 and 4 (Week 3: SAT-WL p<0.001, SAT-WMT p<0.01, Week 4: SAT-WL p<0.001, SAT-WMT p=0.01). No differences were observed between the WMT and WL at any of the time points. Analyses were carried out for each of the 8 areas of interest separately, but for brevity are not reported in full here. All revealed an essentially similar pattern as the combined results with the critical intervention-by-time interaction reaching statistical significance in all cases. In summary, despite the absence of training effects on the standardised cognitive measures reported above, there was a developing pattern over the 4-weeks in which participants completing SAT, relative to the other conditions, perceived their memory, concentration, motivation etc. as improving.

**Figure 4:**
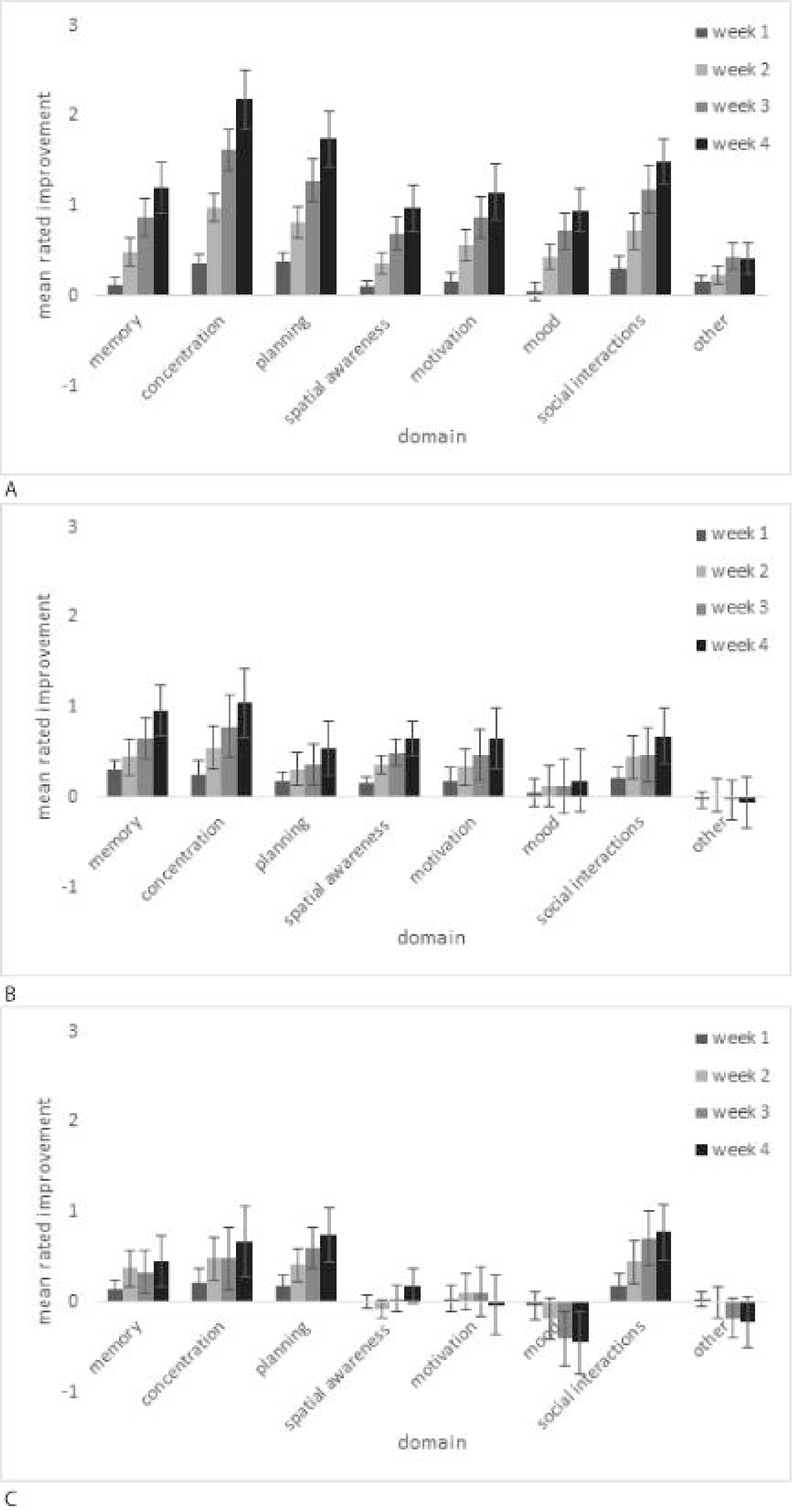
Mean ±S.E. Patient rating of change in symptoms compared to the previous week across 8 key areas, for A. SAT, B. WMT and C. WL.

In our previous study focusing on stroke survivors with likely attentional impairments (Peers et al., 2020), self-reported improvements in functioning were related to improvements in spatial awareness, but not to changes in working memory performance. Here, we again used regression to investigate this issue by examining the degree to which improvement in subjective ratings during the intervention ‘predicted’ post-intervention spatial bias scores, relative to pre-intervention spatial bias scores. Conducting this for each group separately revealed, for the SAT group, post-intervention spatial bias scores were significantly predicted (R^2^= 0.67, F(2,38)=36.49, p<0.001) by ‘pre-intervention’ score and cumulative self-rated improvements at week 4. Unsurprisingly, ‘pre-intervention’ score (β=0.83, p<0.001) was in itself a significant predictor. Consistent with the previous finding, however, and despite the lack of general change in spatial bias highlighted above, improvement in self-ratings showed a trend towards statistical significance in predicting post-intervention bias (β=-0.18, p=0.07) that was absent in the WMT (β=0.07, p=0.53) and WL (β=-0.06, p=0.55) groups. To examine whether improved spatial awareness was associated with self-reported improvements in specific areas (memory, concentration etc.) regressions were carried out separately for each. In both WMT and WL groups, no area significantly predicted post-intervention spatial bias, however for the SAT group, cumulative improvements in planning (β=-0.26, p<0.01) and motivation (β=-0.20, p=0.05) were found to significantly predict post-intervention bias. In summary, although there were no significant overall group changes in spatial bias in any intervention, once again self-reported improvements in cognition and mood were significantly predicted associated with changes in spatial awareness only for those participants in the SAT condition.

Participants completed the European Brain Injury Questionnaire (EBIQ) before and after the intervention. They also, as highlighted above, provided brief ratings of change from the previous week in 8 areas of function in the weekly phone call during the intervention. If the weekly measure of changes in functioning reflect a global changes in symptom severity, it might be expected that the cumulative improvements by week 4 would be a significant predictor of EBIQ core symptoms post training. For the SAT group post-intervention EBIQ core symptoms were significantly predicted by pre-intervention EBIQ core symptoms and cumulative self-rated improvement over the 4 weeks (R^2^= 0.71, F(2,38)=45.03, p<0.001). Both pre-intervention score (β=0.86, p<0.001) and self-rated improvement (β=-0.20, p<0.05) were significant predictors. Self-reported improvement was not found to be a significant predictor of post-intervention EBIQ core symptoms in the WMT group (β=-0.11, p=0.10), though there was a trend towards significance in the WL group (β=-0.22, p=0.08).

#### Who might benefit from training?

To examine whether there were factors that could predict which participants would benefit most strongly from training, a regression analysis was carried out investigating whether the cumulative improvements in self-reported function could be predicted from certain baseline characteristics. These were selected to cover a range of areas including demographics (age, sex and lesion location (left/right/bilateral), fluid intelligence (Cattell Culture Fair), stroke symptom severity (EBIQ core symptoms), intervention (WMT, SAT), and cognitive factors previously associated with the strength of training improvements (TVA absolute spatial bias, K’ variability and AWMA dot matrix; Peers et al., 2020). Together, these factors were shown to significantly predict improvement in self-reported everyday function (R^2^= 0.29, F(10,95)=3.44, p=0.001). Three factors significantly predicted reported improvements in everyday function. These were, being assigned to SAT (β=0.44, p<0.001), having a larger spatial bias at baseline (β=0.28, p=0.006) and having lower levels of baseline K’ variability (β=-0.24, p=0.028).

### 3-month follow-up

Regression analyses examining the influence of pre-intervention performance and intervention type on 3-month follow-up scores are presented in Table 4 below. Given that group level differences were not seen immediately after the interventions, no significant group level effects were expected or observed.

**Table 4.**
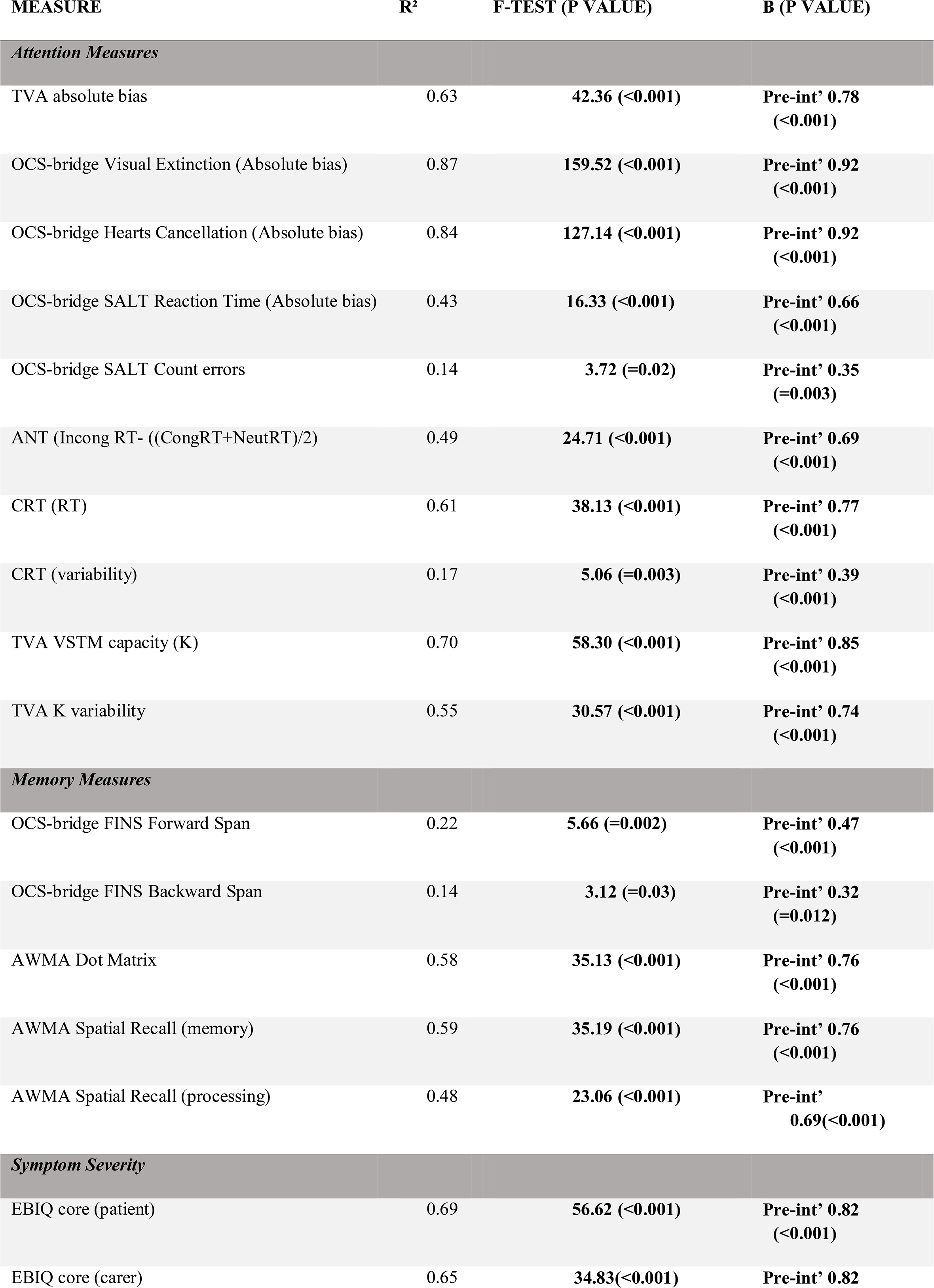
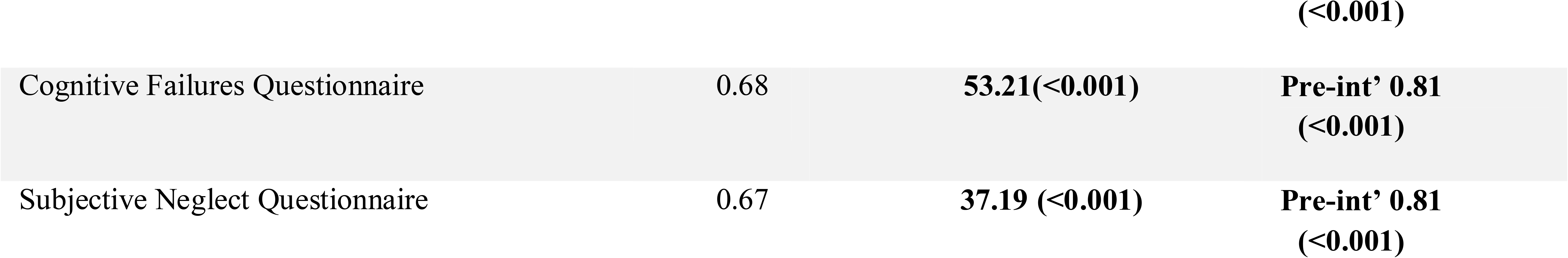
The results of the regression analyses at 3 months post training. The table provides the variance (R^2^) in post-intervention scores that can be predicted by pre-intervention performance score and intervention group, the F-test and *p* value, as well as the standardised β coefficients (and p-values) of significant predictors. Statistically significant results (P<-0.05) are in bold. Int’ = intervention

## Discussion

In a previous study with stroke survivors (Peers et al., 2020) we found encouraging indications that 4 weeks of daily home-based attention training was linked with significant improvements in objectively assessed attention and spatial bias, and in participants’ self-ratings of cognitive and mood function. The results also showed that the training was generally feasible and acceptable. That study had a relatively small group of participants (20), recruited from a panel of keen research volunteers selected on the basis of likely attention impairments. The main aim of the current study was to examine whether these benefits would generalise to a larger group (N = 80) referred by clinicians solely on the basis of being a stroke survivor rather than having particular cognitive problems.

The results here confirmed that the online training could be effectively self-administered in the home in most cases, and that the vast majority of participants had good compliance in completing the recommended training sessions. In the two training conditions, participants showed the expected improvements in performance on the training tasks that, as with our previous report, appeared to level off between 15 and 20 days of training. This high level of compliance (92% of SAT participants and 82% of WMT participants completed the study) and training improvement provide a good basis to judge whether benefits generalised to our primary outcome measure, the TVA measure of spatial bias. In this respect, there was *no* group level evidence to suggest that SAT or WMT were superior to simply being allocated to a waitlist over the same period. This was also the case across all our performance-based measures. There was, in short, no compelling evidence on standardised cognitive assessments of general benefit due to attention or working memory training in this sample.

In contrast, significant differences between the conditions were observed in terms of participants’ weekly self-reports of improvements in areas such as memory, concentration, planning, spatial awareness, motivation and mood. These were greater in the SAT than the WMT and WL conditions. Additionally, post-study interviews revealed that the vast majority of participants viewed the training as manageable (100% for SAT, 90% WMT) and the majority (76% for SAT, 66% for WMT) felt that the training had helped them. If Patient Reported Outcomes (PROs) are considered the best measure of an intervention, then there are grounds to generally recommend training, and specifically SAT, to people who have experienced a stroke. One important caveat in this respect is that, whilst the post-study interviews were carried out by a researcher who was blind to training condition, for practical reasons (e.g. having to solve technical issues), the researcher conducting the weekly interviews was not. It is perhaps significant that the degree to which participants perceived positive improvements was related to post-training spatial bias scores, i.e. that the participants were possibly picking up on changes to which the other performance-based cognitive tests were insensitive. This fits well with our previous findings (Peers et al., 2020) in which changes in spatial bias (as a result of training) significantly predicted more general changes in EBIQ core symptoms.

We will return to the difference between performance-assessed and self-reported changes in function shortly. First, it is important to address the disparity between the results on our primary outcome standardised test measures between this and the previous study (Peers et al., 2020). Certainly, the current study was better powered. There is abundant evidence that low powered studies, cumulatively, tend to exaggerate the effectiveness of interventions. The current result may therefore be a useful correction or help in specifying more precisely for whom the intervention may be most effective. As discussed, unlike the previous study, participants here were not selected on the basis of having particular problems in attention. As illustrated in the comparison of the entire sample’s performance at baseline against normative data from the general population, many of the participants here had very well-preserved cognitive abilities. These included on challenging timed measures of fluid intelligence (where fewer than 10% were formally ‘impaired’) and the ability to hold a sequence of digits in mind and repeat them in reverse order (where only 2% were formally ‘impaired’). A number of studies have addressed whether intensive, computerised ‘brain training’ is of significant benefit in the general population as assessed on standardised cognitive tasks. Whilst some have claimed there are indeed gains (e.g. Jaeggi et al., 2008; Penner et al., 2012) a number of authors have argued that these are quite narrowly restricted to tasks similar to those practised in the training (so called ‘near transfer’ effects) and that evidence for ‘far transfer’ is scant (e.g. Reddick et al., 2012; Shipstead et al., 2012; Owen et al., 2010). With due caveats about the range of functions that we measured and the type and ‘dose’ of training applied, the current study is consistent with the latter camp in suggesting no convincing ‘far transfer’ from daily cognitive training to standardised cognitive tasks in an unselected community stroke sample with many well-preserved cognitive abilities.

In terms of spatial bias, which of necessity (on the basis of Peers et al., 2020) formed our primary outcome measure, only approximately 30% of the current sample’s performance was beyond two standard deviations of the general population mean. In short, either of the training conditions would had to have had a very considerable impact to produce disproportionate improvements on a measure in which approximately 70% of the sample were unimpaired. This raises the question of whether there was any evidence that participants in the current study, who most closely resembled those who took part in Peers et al. (2020), may have shown similar gains? Whilst we were underpowered to formally test effects in the sub-group of patients who were impaired at the outset, a regression analysis indicated that, indeed, those participants with initially the largest spatial biases and greatest variability in performance during the course of the TVA task, *who were allocated to SAT,* were those who reported the greatest cumulative benefits. This, alongside the previous findings of changes in spatial bias following training predicting changes in self-reported core symptoms (Peers et al., 2020), the current ones provide more evidence in favour of the notion that improvements in spatial awareness, brought about by training, may have positive impacts on daily functioning in individuals with spatial awareness difficulties.

As discussed, there has been debate about the most relevant outcome measures, including in stroke (e.g. Price-Haywood et al., 2019). Here, our primary outcome point was a measure of spatial bias. There are good reasons to use highly standardised and sensitive cognitive tests, particularly in aspects of function such as spatial bias where people’s insight is known to be limited (e.g. Langer & Bogousslavsky, 2020). In addition, the presence of persisting spatial bias has been linked with increased difficulties in everyday activities (Jehkonen, Laihosalo, & Keetuenen, 2006; Katz, Hartman-Maeir, Ring, & Sorojer, 1999). The reasonable assumption is that changes on a challenging measure will reflect an improvement in capacity that, without necessarily being aware of it, will also have benefit for participants’ everyday lives. Performance-based, standardised tasks can also be considered as *relatively* independent of participants’ expectations, views about a research study and so on. Patient Reported Outcomes (PROs) are *“any report of the status of a patient’s health condition that comes directly from the patient, without interpretation of the patient’s response by a clinician or anyone else”* (U.S. Department of Health and Human Services FDA Center for Drug Evaluation and Research et al., 2006). There are many outcomes that can only really be judged based on patient report (e.g. pain, mood). In addition, it can be argued that, even when there are objective measures of function (e.g. walking), if an intervention designed to improve it is not reflected in patient-perceived measures of community mobility, quality of life, satisfaction etc., it has not been ‘effective’. An issue with PROs is, of course, that they are relatively more likely to be influenced by expectations and bias. It was for this reason that we compared SAT with a very similar home-training package (WMT) that was of exactly the same duration and on which participants were also likely to observe day-by-day improvements in their performance. In this context, we can be relatively sure that the statistically significant superiority of SAT on PROs does not simply reflect being enrolled in a study, beliefs about the effects of daily cognitive exercise etc. But, in the absence of detectable change across a range of standardised cognitive measures, what can explain the SAT benefit? One possibility is that it did produce greater improvements in cognitive functions to which the participants were sensitive but were missed by our measures. This is not implausible.

Even with an extensive testing battery we are likely to miss many aspects of cognitive function and have tests which are not sufficiently sensitive in the relevant range. It is of note, however, that the PROs lacked specificity; whatever participants were reporting on had apparently had sufficiently diverse influence to affect planning, motivation and mood. Another possibility is that the attribution of greater gains stemmed from participants enjoying SAT more that WMT, which was certainly the case according to their reports; that this enjoyment cast a ‘warm glow’ benefitting reports of cognitive and other gains. Of course, the corollary could also be true; that rated enjoyment was greater because of the perceived cognitive gains. A further possibility in accounting for SAT superiority in PROs is that this training afforded a greater sense of progress. Level of performance in SAT was primarily related to reaction time, which can decrease smoothly with practice. In the WMT, in contrast, level was determined by span length and considerably greater changes in function may be needed to increase span by one item compared with reaction time changes. Whether any, or a combination of, the above accounts is relevant cannot be determined from our current data.

In summary, here we examined whether previously reported benefits of online attention training would generalise to a larger sample of stroke survivors who were not selected on the basis of having likely attentional impairment. As a group they had relatively well-preserved cognitive abilities. They completed 4 weeks of daily online cognitive training in one of two training conditions. Compliance, acceptability and management of the technical demands of the training were high in both conditions and participants showed the expected gains in performance on the trained tasks. Despite this, and contrary to expectations, neither condition was associated with any significant gains on our primary outcome measure of spatial bias, nor any other standardised blind-assessed cognitive test secondary outcome measures, when compared with a waitlist condition. In contrast, patient reported outcomes in areas such as concentration, planning, spatial awareness, motivation and mood did improve, and did so significantly more in the Attention Training, than in the Working Memory Training condition, and also relative to the Waitlist control. With due caveats about researcher blinding in the collection of PROs and whether these gains stemmed from greater enjoyment of the Attention Training, or the possibly greater ease with which improvements are tracked in this condition, the results suggest that Attention Training may have greater influence on PROs than on standardised tests. Whether such training can be recommended for stroke survivors irrespective of cognitive impairment depends on whether objective performance on standardised cognitive tests or subjectively-reported PROs such as mood and motivation are the preferred outcome measures. Further work is required to establish whether the standardised cognitive measure improvements, previously observed in participants with likely attention impairment, replicate in a more similar sample.

## Data Availability

Data will be made open upon acceptance in a journal

## Acknowledgements

This work could not have been carried out without the willingness and ecort of our patients and their families.

## Data Availability

The data are freely available at

## ORCID

Polly V Peers http://orcid.org/0000-0003-4470-9508

