## Supplementary Materials for "A randomised control trial of the effects of home-based online attention training and working memory training on cognition and everyday function in a community stroke sample"

Examples of the training tasks

Working memory training comprised four tasks, a visual and verbal version of both simple span and n-back tasks. In the simple span tasks participants were given serial presentations of either a sequence of (A) locations (spatial task) or digits (verbal task). At the end of the presentation the participant had to indicate, using a mouse, the order of the sequence. Feedback was given after each response. Sequences increased or decreased in length depending upon the participant’s performance over preceding trials. In the n-back tasks participants saw a continuous stream of either locations (spatial task) or letters (verbal task). They were asked to respond when they saw a location or letter that matched the one they had seen ‘n’ items previously. Feedback was given after each response and ‘n’ was increased or decreased based on the accuracy of previous responses.

A

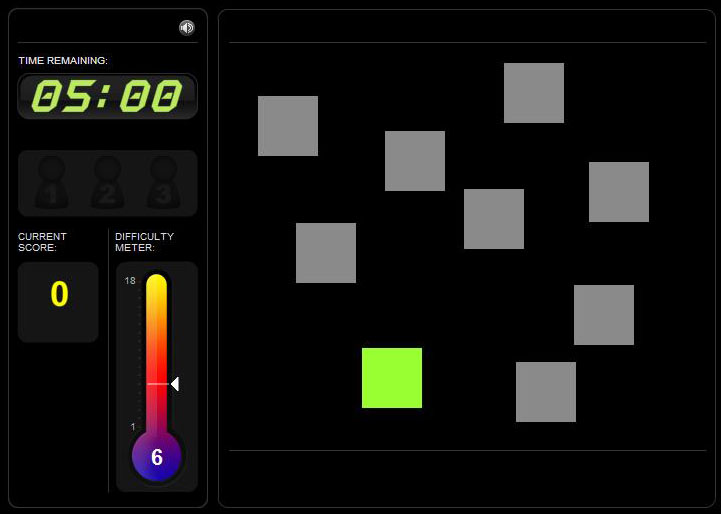

Selective attention training comprised five tasks which were designed to mould attentional selection. The alien task (A) required participants to decide whether an exact match to the alien in the green box was present elsewhere on the screen. Difficulty, based on the number of aliens on the screen, their similarity to one another, and the length of time available to make the decision, was manipulated on the basis of performance over preceding trials. As with all other tasks feedback was provided after each trial. The button sorting task (B) required participants to sort ‘buttons’ based on changing rules of colour or shape. Difficulty was manipulated, dependent on performance, by the frequency of the rule shifts, the congruency of the stimuli and exemplar on the non-target dimension, and the similarity of the target stimulus to the exemplars. For example, in more difficult levels the target stimulus was morphed between the two exemplar characteristics and the participant was requested to sort the button to the exemplar that was most similar to target stimulus. The visual search task (C) required participants to remember physical attributes (size, shape, colour, pattern and orientation) of a single target stimulus and then search for it in a subsequent array of shapes. Difficulty was varied, dependent upon performance on previous trials, by increasing the number of stimuli to search, and how similar these stimuli are the target. The jigsaw task (D) required participants to indicate whether a target jigsaw at the top of the screen could be made from pieces presented at the bottom of the screen. Here difficulty was manipulated by increasing the number of pieces in the jigsaw, time limits and making the discriminations more difficult by rotating pieces. The final task, the rotations task (E) required participants to decide whether two grids, placed in different orientations, would be the same as each other (if in the same orientation) or not. Difficulty here was manipulated by the number of cells in the grid that were filled and the amount of time individuals had to make decisions.

A

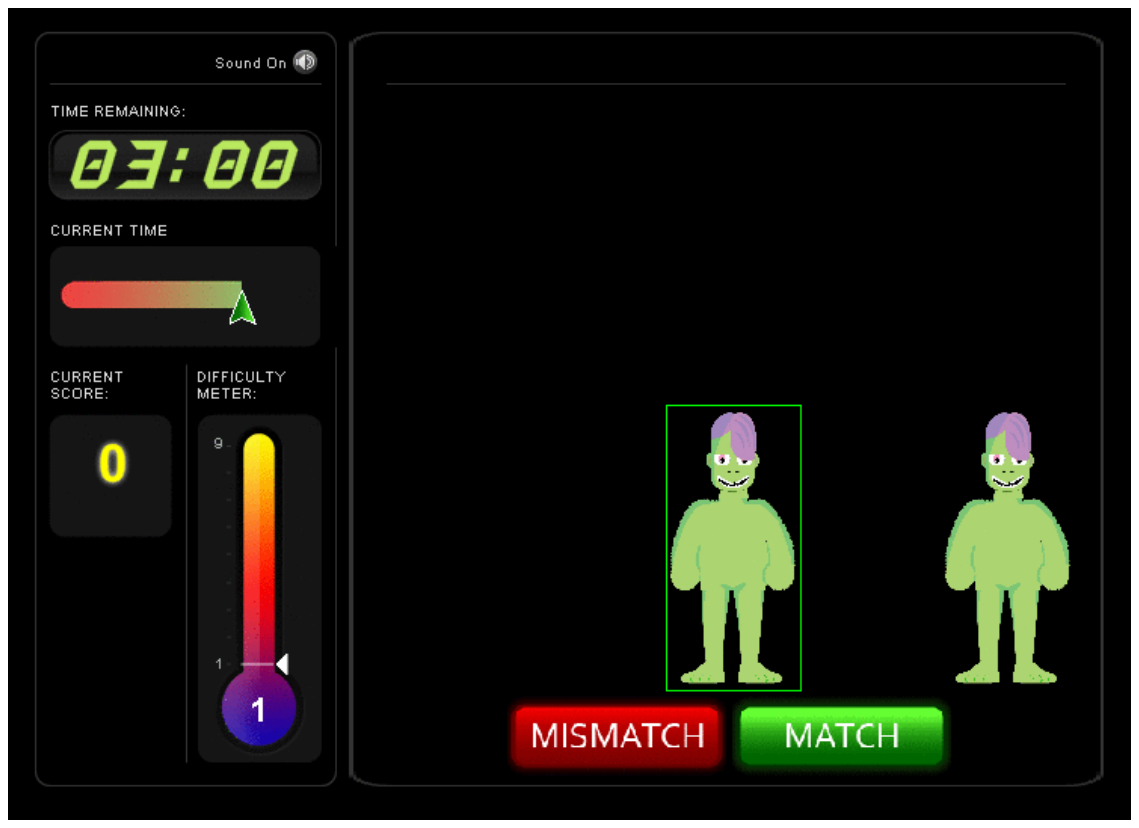

B.

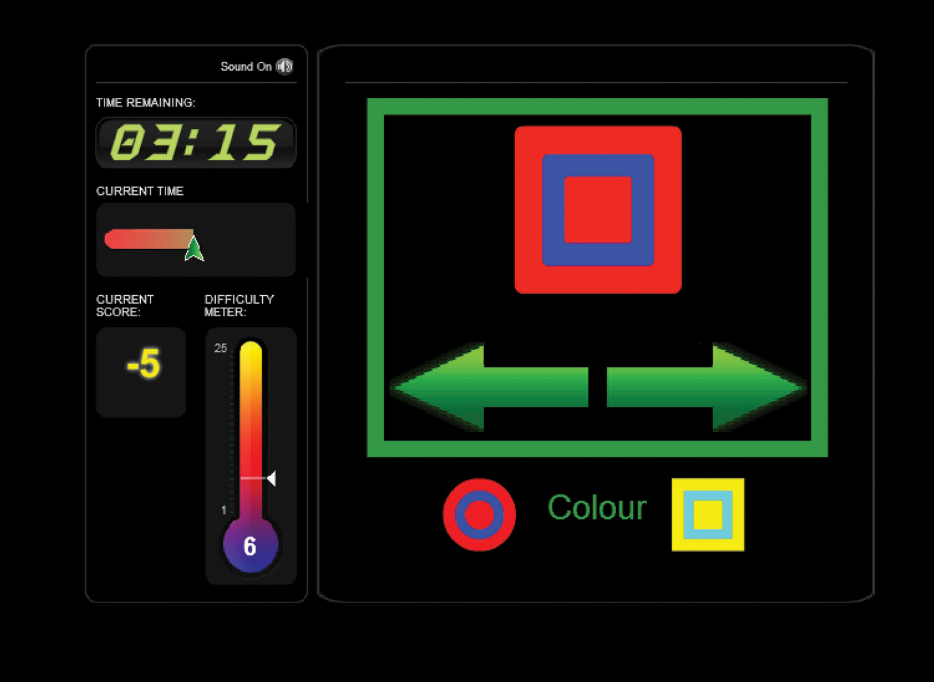

C

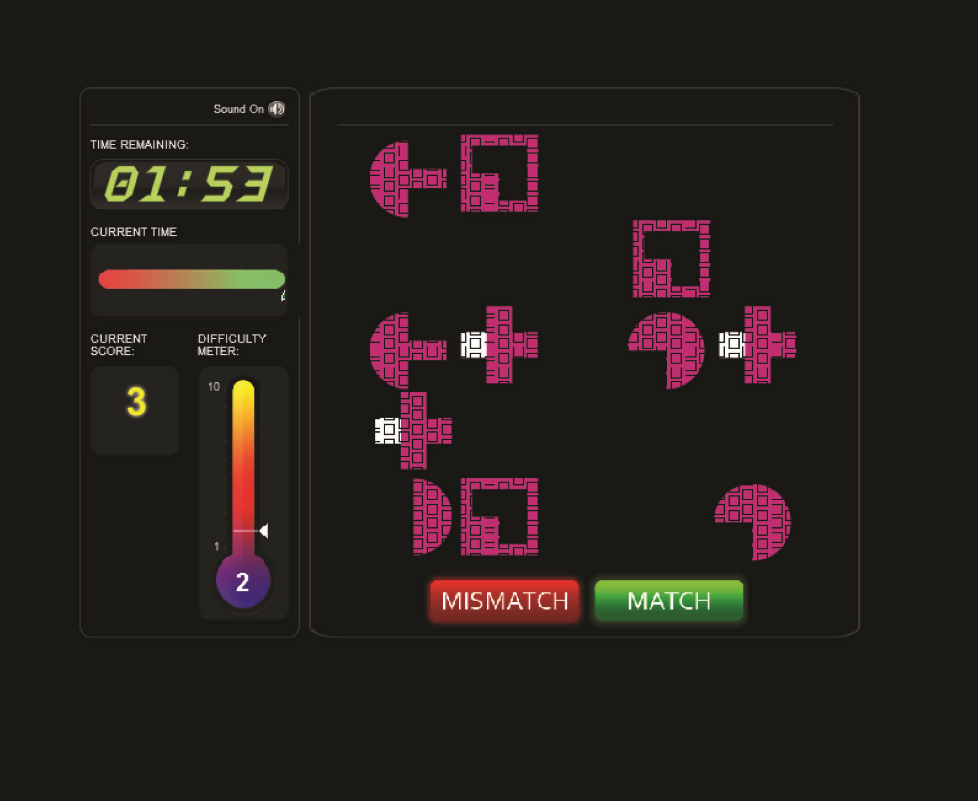

D

.
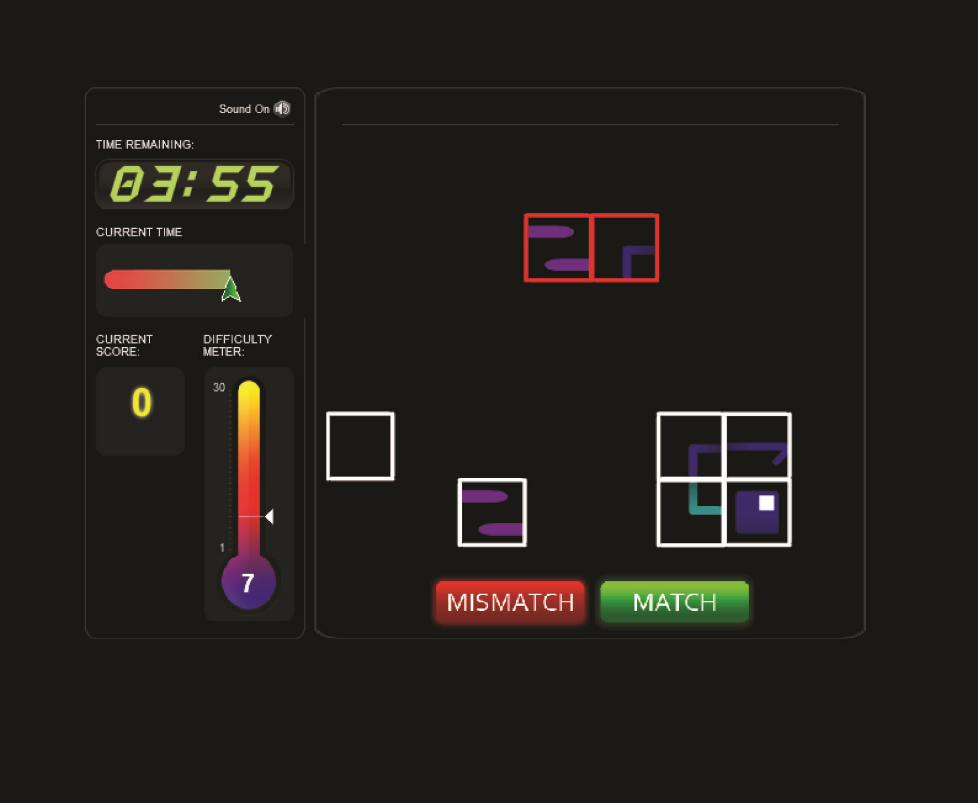

E

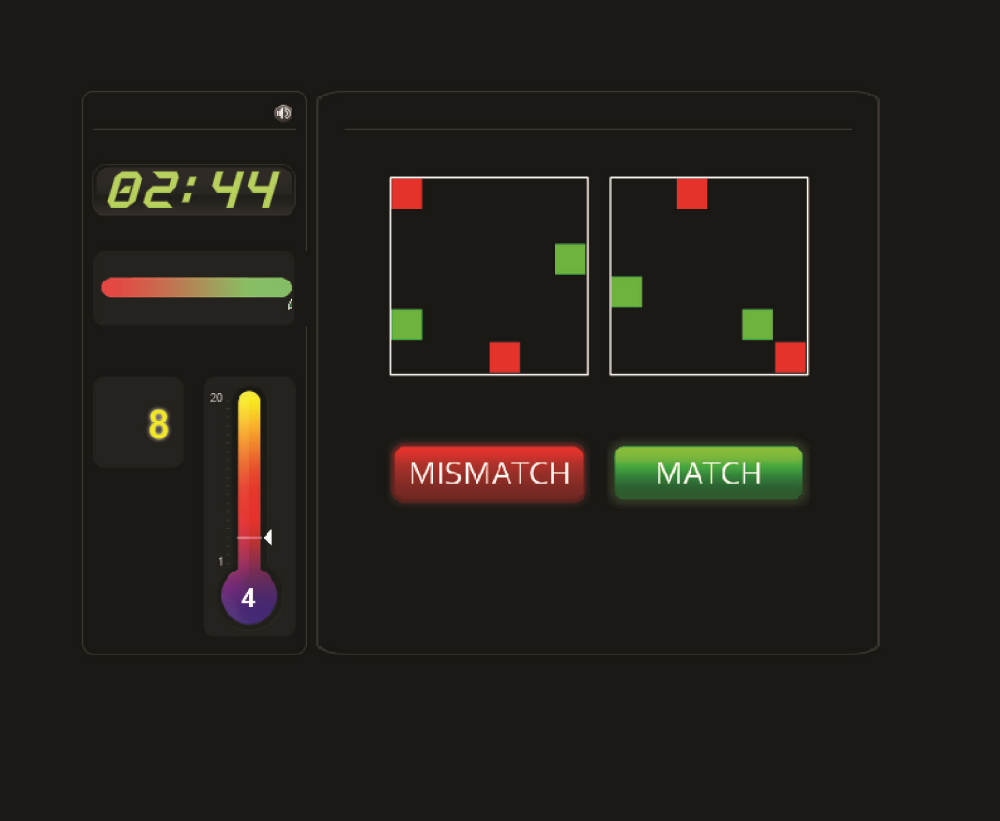

Weekly Telephone Questionnaire

**Weekly telephone questionnaire**

1. How many days have you been able to complete training in the last week?

**0	1	2	3	4	5	6 +**

2. What sort of time are you training?
1. **Morning	2. Afternoon	3. Evening	4. Night**

**5. Different each day**

3. Do you feel that you are making progress?

**Yes	No**

4. Do you see any benefits to your training?

**Yes	No**

5. Do you feel that there are any barriers to you completing the tasks?

**Yes	No**

1. **Motivation	4. Time	7. Effort**
2. **Mood	5. Technology	8. Physical**
3. **Organisation	6. Memory	9. Attention**
6. (If **“Yes”**) What can we do to overcome these barriers?

**“On a scale of 1-10…”**

7. How helpful do you feel the training is?

**1	2	3	4	5	6	7	8	9	10**

8. How enjoyable do you find the training?

**1	2	3	4	5	6	7	8	9	10**

**9.** Have you been able to transfer any of the skills you have used in training to help in your everyday life?

**Yes	No**

**10.** In the last week have you noticed any change in the following areas:

|  | **A lot worse**  **(-2)** | **A bit worse**  **(-1­)** | **No change**  **(0)** | **Improved a bit**  **(+1)** | **Improved a lot**  **(+2)** |
| --- | --- | --- | --- | --- | --- |
| **Your memory** |  |  |  |  |  |
| **Your concentration** |  |  |  |  |  |
| **Your ability to plan/ organise yourself** |  |  |  |  |  |
| **Your spatial awareness** |  |  |  |  |  |
| **Your motivation** |  |  |  |  |  |
| **Your mood** |  |  |  |  |  |
| **Your number of social interactions** |  |  |  |  |  |
| **Or something else** |  |  |  |  |  |

End of Study Interview Questions

**Post Training Questionnaire**

Thank you for taking the time to complete this questionnaire hopefully your responses will help us to develop a more user friendly training program.

***About you***

When did you have your brain injury?

My recovery from my brain injury has been….

Excellent	Very Good	Good	OK	Poor	Very Poor

Compared to before my brain injury I would say I am…

Completely	Not at all

100%	90%	80%	70%	60%	50%	40%	30%	20%	10%	0%

back to normal.

As a result of my brain injury I still have difficulties with (circle all appropriate)….

Movements	My memory

Organization	Impulsivity

Speech	Spatial awareness

Anxiety	Understanding other people

Concentration	Reading

Anger	Short term memory

Vision	Writing

Keeping more than one thing in mind	Apathy

Frustration	My emotions

Remembering things from long ago

Other (please state)

Which of these (if any) is most problematic for you?

***About the training program***

Did you like the idea of a home based skills training program?

yes/ no

Did having to use the computer/internet put you off?

yes/ no

Did you have technical difficulties during the training period?

yes/no

if yes were these due to:	Poor internet connectivity	Inability to understand the tasks

Glitches in the program	Difficulty working your computer

Difficulty seeing/ interacting with the computer	Other

Did other issues make it difficult for you to complete the training?

yes/no

if yes were these due to :	Personal issues	Time

Ability to organise myself	Lack of motivation

Finding a time when it was calm/ quiet	Other

Have you found any strategies helpful in completing your training?

yes/no

If yes what were these.

Did you find the tasks interesting/ engaging?

yes/no

Do you think the 20-30mins of training per day for a month was manageable?

yes/ no/ not sure

Do you think that regular contact with the research team by telephone and/or email was helpful?

yes/no /not sure

Was the frequency of that contact

too infrequent/ about right/ too often?

Do you feel the training helped you?

Yes/ not sure/ no

If you felt it was beneficial how do you think it helped you?

Do you feel there are any improvements we could make? Please continue overleaf if necessary.
